# Pediatric critical COVID-19 and mortality in a multinational cohort

**DOI:** 10.1101/2021.08.20.21262122

**Authors:** Sebastian Gonzalez-Dambrauskas, Pablo Vasquez-Hoyos, Anna Camporesi, Edwin Mauricio Cantillano, Samantha Dallefeld, Jesus Dominguez-Rojas, Conall Francoeur, Anar Gurbanov, Liliana Mazzillo Vega, Steven Shein, Adriana Yock-Corrales, Todd Karsies, on behalf of Critical Coronavirus and Kids Epidemiological (CAKE) Study Investigators

**Author notes:** **Address correspondence to:** Todd Karsies, Division of Critical Care Medicine, Nationwide Children’s Hospital, 700 Children’s Drive, Columbus, Ohio 43205. CAKE Study collaborators listed in an online supplement. **Clinical Trial Registration:** Not applicable. **Contributor’s Statement Page** **Authors’ Contributions:** Drs González-Dambrauskas and Vásquez-Hoyos conceptualized and designed the study, participated in data collection, had full access to all the data in the study and take responsibility for the integrity of the data and the accuracy of the data analysis, helped to draft the initial manuscript, and reviewed and revised the manuscript. Dr. Karsies conceptualized and designed the study, coordinated and supervised all aspects of the study, participated in data collection, performed initial data analysis, had full access to all the data in the study and take responsibility for the integrity of the data and the accuracy of the data analysis, helped to draft the initial manuscript, and reviewed and revised the manuscript. Drs. Camporesi, Cantillano, Dallefeld, Dominguez-Rojas, Francoeur, Gurbanov, Mazzillo Vega, Shein, and Yock-Corrales all participated in data collection, reviewed and revised the manuscript, and critically reviewed the manuscript for important intellectual content. All authors approved the final manuscript as submitted and agree to be accountable for all aspects of the work.

## Abstract

**Objectives:** To understand the international epidemiology of critical pediatric COVID-19 and compare presentation, treatments, and outcomes of younger (<2 years) and older (>2 years) children.

**Design:** Prospective, observational study from April 1 to December 31, 2020

**Setting:** International multicenter study from 55 sites from North America, Latin America, and Europe.

**Participants:** Patients <19 years old hospitalized with critical COVID-19

**Interventions:** none

**Main outcomes measured:** Clinical course, treatments, and outcomes were compared between younger and older children. Multivariable logistic regression was used to calculate adjusted odds ratios (aOR) for hospital mortality.

**Results:** 557 subjects (median age, 8 years; 24% <2 years) were enrolled from 55 sites (63% Latin American). Half had comorbidities. Younger children had more respiratory findings (56% vs 44%), viral pneumonia (45% vs 29%), and treatment with invasive ventilation (50% vs 37). Gastrointestinal (28% vs 69%) or mucocutaneous (16% vs 44%) findings, vasopressor requirement (44% vs 60%), and MIS-C (15% vs 40%) were less common in younger children. Hospital mortality was 10% overall but 15% in younger children (odds ratio 1.89 [95%CI 1.05-3.39]). When adjusted for age, sex, region, and illness severity, mortality-associated factors included cardiac (aOR 2.6; 95%CI 1.07-6.31) or pulmonary comorbidities (aOR 4.4; 95%CI 1.68-11.5), admission hypoxemia (aOR 2.33; 95%CI 1.24-4.37), and lower respiratory symptoms (aOR 2.83; 95%CI 1.49-5.39). Gastrointestinal (aOR 0.49; 95%CI 0.26-0.92) or mucocutaneous symptoms (aOR 0.31; 95%CI 0.15-0.64), treatment with intravenous immune globulin (aOR 0.33; 95%CI 0.17-0.65), and MIS-C (aOR 0.26; 95%CI 0.11-0.64) were associated with lower mortality.

**Conclusions:** We identified age-related differences in presentation and mortality for critical pediatric COVID-19 that should prompt more attention to improving management in younger children, especially in developing countries.

**Table of Contents Summary:** This is a multinational study describing critical pediatric COVID-19 clinical spectrum and related mortality in high and low-middle income countries during 2020.

**What’s known on this subject:** Pediatric critical illness due to COVID-19 is uncommon and have lower mortality compared to adults when hospitalized. While larger cohorts are from high-income countries (HICs), studies including data from low-middle-income countries (LMICs) remain scarce.

**What this study adds:** In our multinational cohort of critical pediatric COVID-19, we identified higher mortality than previously reported and age-related disease patterns. Children <2 years old had more respiratory disease and higher mortality, and older children had more non-pulmonary disease and better outcomes.

## INTRODUCTION

Evidence from the first year of the coronavirus disease 2019 (COVID-19) pandemic indicates that affected children typically develop mild, often asymptomatic, disease and are less likely to require hospitalization or intensive care unit (ICU) admission compared to adults ^1 2^. However, when children do require hospitalization, up to one-third require ICU admission ^2 3^. Few studies focus on pediatric critical COVID-19, and most larger studies are from high-income countries (HICs) ^4–7^. Studies examining pediatric ICU (PICU) admissions from low-middle-income countries (LMICs) are scarce, and the main focus has been multisystem inflammatory syndrome in children (MIS-C) ^8–10^.

In addition to a less severe presentation, children have lower mortality compared to adults, even when hospitalized ^1 2 11^. While pediatric COVID-19 mortality rates are low in HICs, they are more concerning in LMICs. The review by Kitano and colleagues found that over 90% of pediatric COVID-19 deaths were from LMICs and that mortality risk was highest in children under 1 year old ^12^. Mortality differences in LMICs can be partly explained by less-developed healthcare infrastructure and lack of resources, but the reasons for age-related differences are unclear. Potential explanations include developmental immune function differences, lack of prior exposure to other coronaviruses, and less frequent development of MIS-C, which might have lower mortality compared to COVID-19-related acute respiratory disease ^13 14^. This mortality risk is further complicated by the fact that young children are currently unlikely to receive SARS-CoV2 vaccination because of lack of regulatory approval for this age group, the perception that young children are unlikely to have severe infections, and public health strategies that prioritize adult vaccination.

To understand the epidemiology and outcomes of critical pediatric COVID-19 in both HICs and LMICs, we designed the Critical Coronavirus And Kids Epidemiology (CAKE) study, which includes PICUs from North, Central, and South America plus Europe. We previously reported a case series with preliminary insights into pediatric critical COVID-19 across these regions ^15^. The objective of this manuscript is to more fully describe the epidemiology, clinical findings, and hospital treatments of critical COVID-19 in children from HICs and LMICs, as well as to identify factors associated with outcomes. Due to suspected differences in both the clinical presentation and outcomes between younger children (<2 years old) and older children (>2 years old) based on our experience with other respiratory viruses (i.e. bronchiolitis) as well as our early experiences with COVID-19, we compared younger and older children under the hypothesis that younger children with critical COVID-19 have a different clinical phenotype and worse outcomes than older children.

## METHODS

### Study design and setting

CAKE is a prospective, observational cohort study examining the epidemiology and outcomes of children hospitalized for severe or critical COVID-19. We used STROBE guidelines when reporting.

### Participants

We enrolled patients <19 years old hospitalized between April 1 and December 31, 2020, with severe or critical confirmed COVID-19 and/or MIS-C. Patients were enrolled if they met the following criteria:

- Laboratory proven evidence of current or prior infection by SARS-CoV2
- COVID-19 related illness as the primary reason for hospitalization And either
- Severe or critical COVID-19 based on our prior definition ^15^ Or
- MIS-C requiring ICU admission, even if not meeting our definition of severe/critical COVID-19.

Laboratory confirmation of SARS-CoV2 infection was done using polymerase chain reaction, antigen, or serologic testing depending on laboratory availability.

### Definitions

Critical COVID-19 was defined as having a positive test for SARS-CoV2 and requiring at least one of following therapies: ICU-level respiratory support including high flow nasal cannula (HFNC), non-invasive ventilation (NIV), or invasive mechanical ventilation (IMV); intravenous vasopressor or inotropic support; or continuous renal replacement therapy (CRRT). Severe COVID-19 included patients with a positive SARS-CoV2 test who did not meet critical COVID-19 criteria but received oxygen support via mask or nasal cannula exceeding the pediatric acute respiratory distress syndrome (ARDS) “at risk” criteria ^16^. MIS-C cases were defined using the Centers for Disease Control and Prevention (CDC) criteria ^17^. Because shortness of breath can be associated with both cardiovascular and respiratory disease processes, we opted to categorize this symptom as “non-specific” rather than “respiratory.”

### Data collection, variables, and outcome measures

Prospective de-identified data were collected using a modification of the case report form developed by the International Severe Acute Respiratory and emerging Infection Consortium (ISARIC). We used REDCap (Research Electronic Data Capture, Vanderbilt University) online database hosted by Nationwide Children’s Hospital (Columbus, Ohio, USA). Collected data included demographics, comorbidities, presenting symptoms, epidemiologic exposures, microbiologic data, ICU and hospital treatments (including organ support and medications), duration of treatments, hospital and PICU length of stay (LOS) and in-hospital mortality. Data on illness progression and severity were collected daily from hospital day 1 until day 14, death, or hospital discharge. Illness severity was estimated using the Pediatric Risk of Mortality III (PRISM III) and Pediatric Logistic Organ Dysfunction 2 (PELOD-2) scores ^18 19^. Diagnoses and complications were based on clinician documentation only. Missing data were completed after the enrolment period through record review by each site investigator. Subjects with incomplete data related to presentation, day 1 treatments, or outcomes were excluded from final analysis.

### Data analysis

Categorical variables were described as frequencies and percentages, and continuous variables expressed as medians and interquartile ranges (IQRs) or ranges. We used Fisher’s exact test to analyze categorical variables and Wilcoxon rank-sum test for continuous variables. We performed univariate and multivariate logistic regression to examine associations with mortality; results from regression models are reported as odds ratios (unadjusted or adjusted) and 95% confidence intervals (CI). Statistical significance was designated as p value □0.05 (2-sided) or a 95% confidence interval that does not include 1. For our multivariable model, we included variables that were statistically significant in univariate logistic regression as well as those demonstrated in prior studies to have an association with mortality. Multivariable models were adjusted for age, sex, region (Latin America vs. non-Latin America), and PRISM III, which were selected *a priori* due to suspicion that they may be associated with outcomes. Statistical analysis was performed using JMP 15 Pro for Mac (SAS Institute, Cary, North Carolina).

### Ethics

This study was approved by the lead site, Nationwide Children’s Institutional Review Board, and each institutional review board at each participating site with a waiver of the need for informed consent (supplemental data, collaborators).

## RESULTS

A total 84 sites screened patients for inclusion, and 590 subjects were enrolled from 55 different sites. After excluding 33 subjects due to incomplete data from presentation, day 1 laboratory values or treatments, or outcomes, there were 557 subjects included for analysis. Of these, 433 met our criteria for critical COVID-19, 76 met our definition for severe COVID-19, and 48 were in the PICU with MIS-C.

### Demographics and Comorbidities

The median age was 8 years with 24% under 2 years old and 61% male (Table 1). Only 43% had a known COVID-19 exposure. There were 352 (63%) from Latin America (Supplemental Table 1). Comorbidities were present in just over half of subjects (Table 1). Most were similar between older and younger children, although younger children were more likely to have a chronic cardiac condition or malnutrition while older children more frequently had obesity or asthma.

**Table 1:** Demographics, comorbidities, and clinical presentation of children with critical COVID-19.

|  | All<br>(n=557)<sup>a</sup> | Age <2 years<br>(n=134)<sup>a</sup> | Age >2 years<br>(n=423)<sup>a</sup> |
| --- | --- | --- | --- |
| **Demographics** |  |  |  |
| Median Age, years | 8 (2, 12.4) | 0.39 (0.1, 0.94) | 10 (6, 13.6) |
| Male Sex | 338 (61%) | 78 (58%) | 260 (61%) |
| **Comorbidities** |  |  |  |
| Cardiac* | 43 (7.7%) | 24 (18%) | 19 (4.4%) |
| Pulmonary (not asthma) | 32 (5.7%) | 9 (6.7%) | 23 (5.4%) |
| Asthma* | 44 (7.9%) | 1 (0.7%) | 43 (10%) |
| Renal | 22 (4%) | 3 (2.2%) | 19 (4.5%) |
| Liver | 9 (1.6%) | 3 (2.2%) | 6 (1.4%) |
| Neurologic | 58 (10%) | 15 (11%) | 43 (10%) |
| Malignancy | 20 (3.6%) | 2 (1.5%) | 18 (4.3%) |
| Hematologic | 18 (3.2%) | 5 (3.7%) | 13 (3.1%) |
| HIV | 2 (0.4%) | 1 (0.7%) | 1 (0.2%) |
| Obesity* | 74 (13%) | 3 (2.2%) | 71 (17%) |
| Diabetes | 11 (2%) | 0 | 11 (2.6%) |
| Rheumatologic | 2 (0.4%) | 1 (0.7%) | 1 (0.2%) |
| Malnutrition* | 36 (6.4%) | 15 (11%) | 21 (5%) |
| >1 Comorbidity | 119 (21%) | 28 (21%) | 91 (22%) |
| None | 272 (49%) | 63 (47%) | 209 (49%) |
| **Clinical Presentation** |  |  |  |
| Known COVID Exposure | 240 (43%) | 50 (38%) | 190 (45%) |
| Day of Illness at Admission** | 4 (2, 6) | 2 (1, 4) | 4 (2, 6) |
| ICU During Hospitalization | 540 (97%) | 131 (98%) | 409 (97%) |
| Ward Admission Pre-ICU | 249 (47%) | 58 (46%) | 191 (48%) |
| Fever (>38.3°C) at Admission* | 112 (21%) | 13 (10%) | 99 (24%) |
| Admission O<sub>2</sub> Saturation <94% | 116/533 (22%) | 32/129 (25%) | 84/404 (21%) |
| Dehydration at Admission | 87/555 (16%) | 15/133 (11%) | 72/422 (17%) |
| Delayed Capillary Refill at Admission | 141/370 (38%) | 36/101 (36%) | 105/269 (39%) |
| PRISM III at ICU Admission** | 5 (2, 10) | 4 (0, 10) | 5 (2, 10) |
| PELOD 2 at ICU Admission | 2 (0, 6) | 2 (0, 6) | 3 (0, 5) |
| **Presenting Symptoms** |  |  |  |
| Lower Respiratory* | 262 (47%) | 75 (56%) | 187 (44%) |
| Upper Respiratory* | 165 (30%) | 50 (37%) | 115 (27%) |
| Systemic* | 469 (84%) | 91 (68%) | 378 (89%) |
| Gastrointestinal* | 327 (59%) | 37 (28%) | 290 (69%) |
| Neurologic* | 214 (38%) | 41 (31%) | 173 (41%) |
| Mucocutaneous* | 208 (37%) | 21 (16%) | 187 (44%) |
| Non-Specific* | 299 (54%) | 58 (43%) | 241 (57%) |

Data are n (%) or median (IQR)
<sup>a</sup>Sample size unless otherwise noted. Lower sample size due to missing data
\*p<0.05 using Fisher's exact test
\*\*p<0.05 using Wilcoxon rank-sum test
Abbreviations: PRISM III = Pediatric Risk of Mortality III; PELOD-2 = Pediatric Logistic Organ Dysfunction 2

### Exposures and Presentation

Initial presentation and clinical findings are noted in Table 1 and Supplemental Table 2. Younger children presented earlier in their illness and were more likely to have respiratory symptoms but less likely to have gastrointestinal or mucocutaneous symptoms. They were also less likely to have a history of fever or documented temperature >38.3°C on admission.

On presentation, older children were more likely to have laboratory abnormalities, including lymphopenia, thrombocytopenia, hyponatremia, and elevated inflammatory markers (procalcitonin, C-reactive protein, and IL-6) (Table 2). Abnormal cardiac labs (brain natriuretic peptide [BNP] and troponin I) as well as elevated d-dimer were more frequently found in older children. While diagnoses and complications were similar between age groups (Table 3; Supplemental Table 3), viral pneumonia, bacteremia and cardiac arrest did occur more frequently in younger children; myocarditis, acute kidney injury, and MIS-C occurred more commonly in older children.

**Table 2:** Laboratory abnormalities identified on hospital day 1. Percentage reported is based on total number of subjects.

|  | All<br>(n=557) | Age <2 years<br>(n=134) | Age >2 years<br>(n=423) |
| --- | --- | --- | --- |
| Leukocyte Count |  |  |  |
| >15000 cells/mL | 117 (21%) | 26 (19%) | 91 (22%) |
| <4000 cells/ mL | 40 (7.2%) | 6 (4.5%) | 34 (8%) |
| Lymphopenia (<1000 cells/mL) * | 144 (26%) | 5 (3.7%) | 139 (33%) |
| Platelets <150000 cells/mL * | 153 (27%) | 18 (13%) | 135 (32%) |
| Haematocrit < 30% | 129 (23%) | 34 (25%) | 95 (22%) |
| ALT > 50 U/L * | 118 (21%) | 17 (13%) | 101 (24%) |
| AST > 50 U/L * | 130 (23%) | 18 (13%) | 112 (26%) |
| Total Bilirubin > 2 mg/dL * | 36 (6.5%) | 14 (10%) | 22 (5.2%) |
| BUN > 20 mg/dL * | 152 (27%) | 16 (12%) | 136 (32%) |
| Sodium < 135 mEq/L * | 143 (26%) | 13 (9.7%) | 130 (31%) |
| Lactate > 4 mmol/L | 51 (9.2%) | 9 (6.7%) | 42 (9.9%) |
| Procalcitonin > 2 ng/mL * | 65 (12%) | 2 (1.5%) | 63 (15%) |
| C-Reactive Protein > 4 mg/dL * | 255 (46%) | 26 (19%) | 229 (54%) |
| D-Dimer |  |  |  |
| >4 ng/mL FEU * | 191 (34%) | 26 (19%) | 165 (39%) |
| >1000 ng/mL FEU * | 109 (20%) | 11 (8.2%) | 98 (23%) |
| BNP |  |  |  |
| >60 pg/mL * | 129 (23%) | 13 (9.7%) | 116 (27%) |
| >1000 pg/mL * | 74 (13%) | 7 (5.2%) | 67 (16%) |
| Troponin I > 0.03 ng/mL * | 125 (22%) | 19 (14%) | 106 (25%) |
| IL-6 >10 pg/mL * | 44 (7.9%) | 3 (2.2%) | 41 (9.7%) |
| Ferritin >1000 mg/L | 65 (12%) | 6 (4.5%) | 59 (14%) |

Data are n (%)
\* p<0.05 using Fisher's exact test
**Abbreviations:** ALT = alanine aminotransferase; AST = aspartate aminotransferase;
BUN = blood urea nitrogen; BNP = brain natriuretic peptide; IL-6 = interleukin 6
**SI Conversion Factors:** To convert ALT or AST, multiply by 0.0167. To convert bilirubin, multiply by 17.1.
To convert BUN, multiply by 0.357. To convert C-reactive protein, multiply by 10
To convert d-dimer, multiply by 0.55.

**Table 3:** Hospital Diagnoses, Complications, and Outcomes.

|  | All<br>(n=557) | Age <2 years<br>(n=134) | Age >2 years<br>(n=423) |
| --- | --- | --- | --- |
| **Diagnoses/Complications** |  |  |  |
| Viral Pneumonia/Pneumonitis* | 183 (33%) | 60 (45%) | 123 (29%) |
| Bacterial Pneumonia | 113 (20%) | 26 (19%) | 87 (21%) |
| ARDS | 157 (28%) | 42 (31%) | 117 (27%) |
| Seizure | 53 (9.5%) | 15 (11%) | 38 (9%) |
| Stroke | 19 (3.4%) | 5 (3.7%) | 14 (3.3%) |
| Heart Failure | 86 (15%) | 16 (12%) | 70 (17%) |
| Myocarditis/Cardiomyopathy** | 101 (18%) | 11 (8.2%) | 90 (21%) |
| Arrhythmia | 39 (7%) | 9 (6.7%) | 30 (7.1%) |
| Cardiac Arrest* | 56 (10%) | 23 (17%) | 33 (7.8%) |
| Bacteremia** | 68 (12%) | 28 (21%) | 40 (9.5%) |
| Coagulopathy | 98 (18%) | 18 (13%) | 80 (19%) |
| Acute Kidney Injury* | 88 (16%) | 13 (9.7%) | 75 (18%) |
| Pancreatitis | 13 (2.3%) | 1 (0.7%) | 12 (2.8%) |
| Acute Hepatic Injury* | 51 (9.2%) | 6 (4.5%) | 45 (11%) |
| MIS-C** | 188 (34%) | 20 (15%) | 168 (40%) |
| **Outcomes** |  |  |  |
| NIV Duration, days | 2 (1, 5) | 2 (1, 6.5) | 2 (1.4, 4) |
| IMV Duration, days | 5 (2.5, 8.5) | 5 (2.4, 9) | 4.4 (2.4, 8.4) |
| Vasopressor Duration, days | 3 (2, 5) | 4 (2, 6) | 3 (2, 5) |
| ICU LOS, days | 5 (3, 9) | 5 (3, 9) | 5 (3, 8) |
| Hospital LOS, days | 9 (6, 15) | 8.4 (5, 16) | 9 (6, 15) |
| Tracheostomy | 7 (1.3%) | 0 | 7 (1.7%) |
| Mortality* | 56 (10.1%) | 20 (15%) | 36 (8.6%) |
| Day of Death | 6 (3, 17) | 5 (3.2, 10.5) | 6 (3, 25) |

Data are n (%) or median (IQR)
\* p<0.05
\*\* p<0.0001
Abbreviations: ARDS = acute respiratory distress syndrome; MIS-C = multisystem inflammatory syndrome in children; NIV = non-invasive ventilation; IMV = invasive mechanical ventilation; ICU = intensive care unit; LOS = length of stay

### Medications and treatments

The most common ICU-level support modalities included vasopressors, IMV, and NIV (Table 4). IMV was more frequent in younger children, while older children were more likely to require vasopressors. Nearly 1/3 of younger children (31%) and 23% of older children required IMV on the first hospital day (p=0.06). In conjunction with more frequent systemic inflammation and concern for MIS-C, older children more frequently received prophylactic anticoagulation, antiplatelet therapy, immunomodulators (primarily intravenous immune globulin [IVIG]), and methylprednisolone. Exchange transfusions, convalescent plasma, and immunomodulators other than IVIG were infrequent in all age groups. Dexamethasone was more common in respiratory illness (38% of those with lower respiratory symptoms versus 20% without lower respiratory symptoms), but use was similar between older and younger children.

**Table 4:**
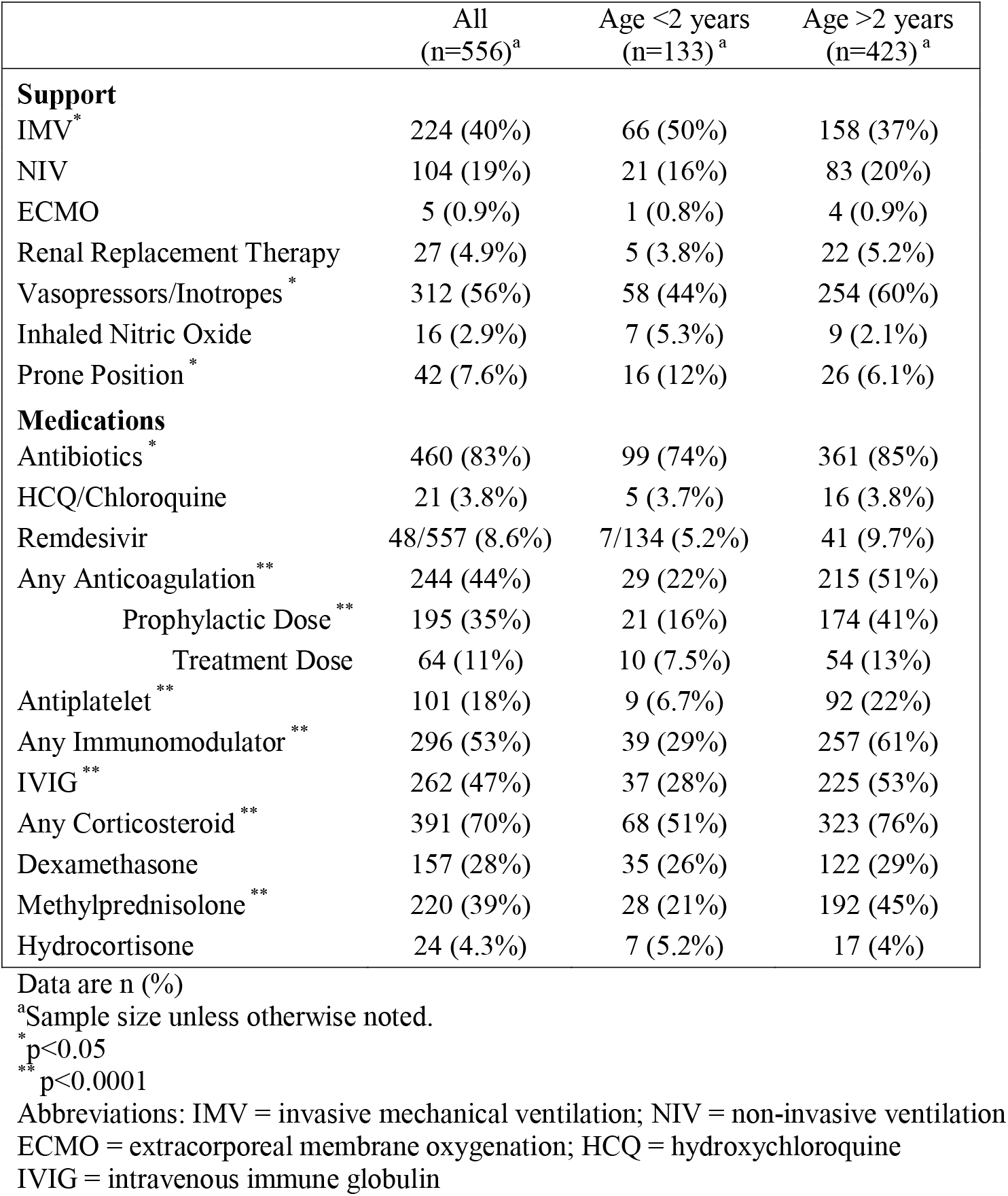
Organ support and medications during hospitalization.

|  | All<br>(n=556)<sup>a</sup> | Age <2 years<br>(n=133)<sup>a</sup> | Age >2 years<br>(n=423)<sup>a</sup> |
| --- | --- | --- | --- |
| **Support** |  |  |  |
| IMV* | 224 (40%) | 66 (50%) | 158 (37%) |
| NIV | 104 (19%) | 21 (16%) | 83 (20%) |
| ECMO | 5 (0.9%) | 1 (0.8%) | 4 (0.9%) |
| Renal Replacement Therapy | 27 (4.9%) | 5 (3.8%) | 22 (5.2%) |
| Vasopressors/Inotropes * | 312 (56%) | 58 (44%) | 254 (60%) |
| Inhaled Nitric Oxide | 16 (2.9%) | 7 (5.3%) | 9 (2.1%) |
| Prone Position * | 42 (7.6%) | 16 (12%) | 26 (6.1%) |
| **Medications** |  |  |  |
| Antibiotics * | 460 (83%) | 99 (74%) | 361 (85%) |
| HCQ/Chloroquine | 21 (3.8%) | 5 (3.7%) | 16 (3.8%) |
| Remdesivir | 48/557 (8.6%) | 7/134 (5.2%) | 41 (9.7%) |
| Any Anticoagulation ** | 244 (44%) | 29 (22%) | 215 (51%) |
| Prophylactic Dose ** | 195 (35%) | 21 (16%) | 174 (41%) |
| Treatment Dose | 64 (11%) | 10 (7.5%) | 54 (13%) |
| Antiplatelet ** | 101 (18%) | 9 (6.7%) | 92 (22%) |
| Any Immunomodulator ** | 296 (53%) | 39 (29%) | 257 (61%) |
| IVIG ** | 262 (47%) | 37 (28%) | 225 (53%) |
| Any Corticosteroid ** | 391 (70%) | 68 (51%) | 323 (76%) |
| Dexamethasone | 157 (28%) | 35 (26%) | 122 (29%) |
| Methylprednisolone ** | 220 (39%) | 28 (21%) | 192 (45%) |
| Hydrocortisone | 24 (4.3%) | 7 (5.2%) | 17 (4%) |

Data are n (%)
<sup>a</sup>Sample size unless otherwise noted.
\*p<0.05
\*\*p<0.0001
Abbreviations: IMV = invasive mechanical ventilation; NIV = non-invasive ventilation
ECMO = extracorporeal membrane oxygenation; HCQ = hydroxychloroquine
IVIG = intravenous immune globulin

### Outcomes

Duration of NIV, IMV, ICU stay, hospitalization, and vasopressor use were similar between age groups. Overall, mortality was 10% (56 subjects) with higher mortality seen in younger children (Table 3). Notably, cardiac arrest was associated with an 85% mortality. Four out of five patients treated with extracorporeal membrane oxygenation (ECMO) survived. We evaluated factors related to mortality using logistic regression with adjustment for age, sex, region, and initial PRISM III score (Figure 1; Supplemental Table 4). Factors associated with higher mortality included cardiopulmonary comorbidities, malignancy, malnutrition, hypoxemia or lower respiratory symptoms at admission, bacteremia, and significant organ dysfunction, such as ARDS, seizures, acute kidney injury, and acute liver injury. Having gastrointestinal or mucocutaneous symptoms, receiving IVIG, and being diagnosed with MIS-C were all associated with lower odds of mortality.

**Figure 1:**
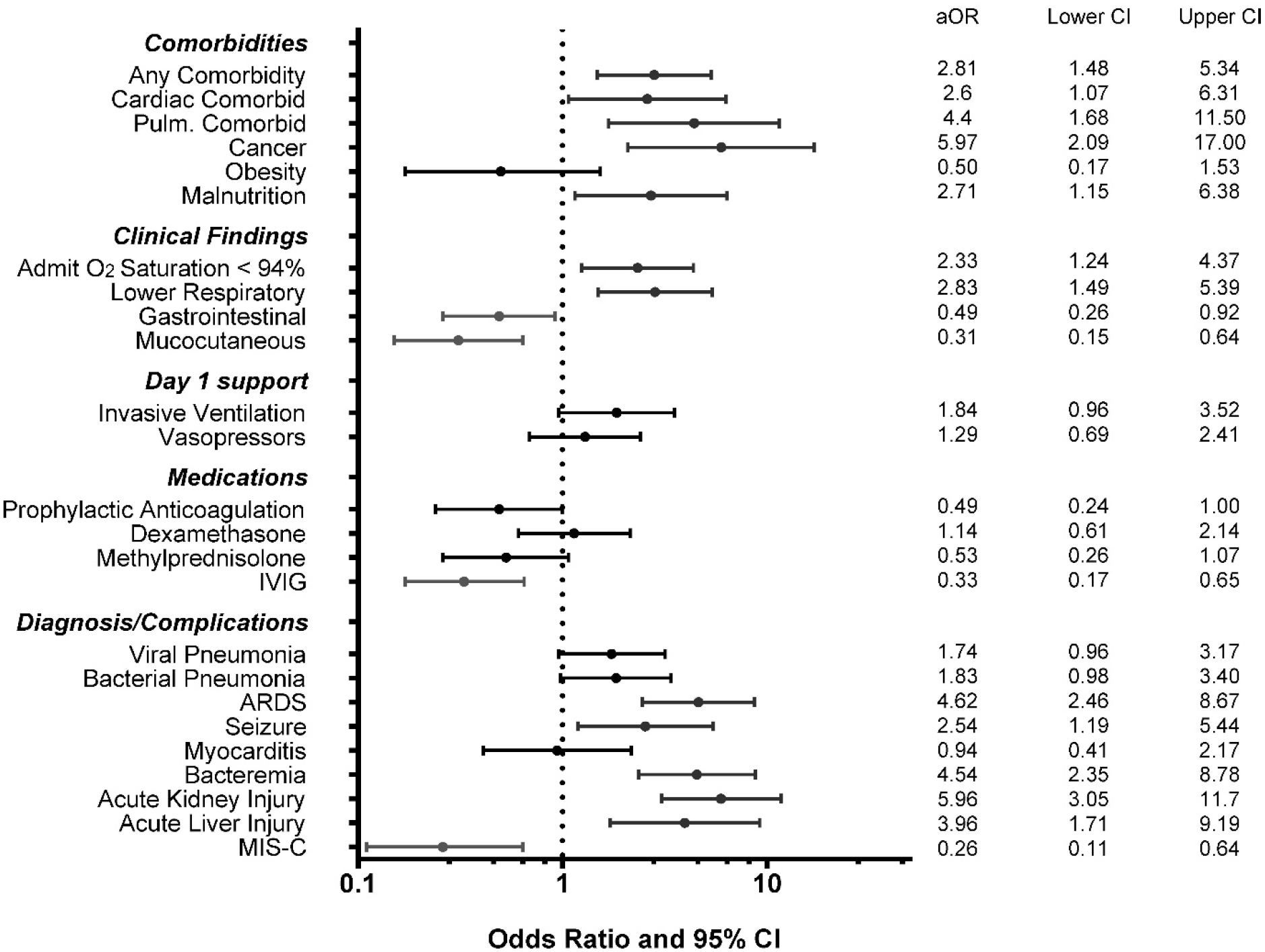
Multivariable Associations with Mortality adjusting for sex, age<2, region, and admission PRISM III. Abbreviations: PRISM III= Pediatric Risk of Mortality III; IVIG = intravenous immune globulin ARDS = acute respiratory distress syndrome; MIS-C = multisystem inflammatory syndrome in children

Because we suspected that clinical factors and treatments may differentially impact mortality in younger children, we explored associations with mortality using univariate logistic regression stratified by age <2 and >2 years old (Table 5). Although most factors significantly associated with mortality in the entire cohort had similar associations in stratified analysis, several had a differential effect depending on age. Prophylactic anticoagulation, IVIG, and methylprednisolone were associated with lower mortality only in older children. Similarly, having a diagnosis of MIS-C or symptoms commonly seen in MIS-C was associated with lower mortality only in older children. Pulmonary involvement, liver injury, or kidney injury were associated with higher mortality regardless of age.

**Table 5:** Univariate associations with mortality plus mortality rate stratified by age. Includes factors significantly associated with mortality in multivariable analysis and other select treatments and diagnoses.

| Variable | <2 years old |  |  | >2 years old |  |  |
| --- | --- | --- | --- | --- | --- | --- |
|  | OR | 95% CI | Mortality | OR | 95% CI | Mortality |
| Admission O<sub>2</sub> Saturation <94% | 2.33 | 0.86, 6.36 | 25% | 2.46 | 1.18, 5.13 | 15% |
| Pulmonary Comorbidity | 1.68 | 0.32, 8.75 | 22% | 4.33 | 1.59, 11.8 | 26% |
| Malnutrition | 3.43 | 1.03, 11.4 | 33% | 3.72 | 1.28, 10.8 | 24% |
| Lower Respiratory Symptoms | 5.57 | 1.55, 20.1 | 23% | 2.72 | 1.32, 5.61 | 13% |
| Gastrointestinal Symptoms | 0.88 | 0.30, 2.63 | 14% | 0.43 | 0.21, 0.85 | 6.3% |
| Mucocutaneous Symptoms | 0.55 | 0.12, 2.57 | 9.5% | 0.33 | 0.15, 0.75 | 4.3% |
| Day 1 Invasive Ventilation | 3.49 | 1.31, 9.31 | 28% | 3.51 | 1.74, 7.07 | 18% |
| Prophylactic Anticoagulation | 0.99 | 0.23, 3.77 | 15% | 0.38 | 0.17, 0.85 | 4.6% |
| IVIG | 1.13 | 0.40, 3.21 | 16% | 0.19 | 0.08, 0.44 | 3.1% |
| Dexamethasone | 0.92 | 0.31, 2.76 | 14% | 1.27 | 0.61, 2.62 | 9.9% |
| Methylprednisolone | 1.30 | 0.43, 3.96 | 18% | 0.26 | 0.11, 0.62 | 3.7% |
| Viral Pneumonia | 4.53 | 1.54, 13.3 | 25% | 1.25 | 0.60, 2.58 | 8.1% |
| ARDS | 7.08 | 2.49, 20.2 | 33% | 6.56 | 3.15, 13.6 | 21% |
| Bacteremia | 5.86 | 2.09, 16.5 | 36% | 6.37 | 2.89, 14.1 | 30% |
| Acute Kidney Injury | 18.2 | 4.76, 69.4 | 67% | 4.42 | 2.17, 9.02 | 21% |
| Acute Liver Injury | 13.9 | 2.35, 82 | 67% | 2.20 | 0.90, 5.37 | 16% |
| MIS-C | 0.99 | 0.26, 3.77 | 15% | 0.12 | 0.04, 0.40 | 1.8% |

IVIG = intravenous immune globulin; ARDS = acute respiratory distress syndrome
MIS-C = multisystem inflammatory syndrome in children

## DISCUSSION

Our study of critical pediatric COVID-19 from HICs and LMICs found distinct age-related disease patterns and outcomes and observed higher mortality than previously reported in large PICU studies ^4 6^. Factors related to mortality included younger age, cardiopulmonary comorbidities, malignancy, malnutrition, and having a primarily respiratory illness (in contrast to MIS-C). Dexamethasone was not associated with improved mortality in any age group, while methylprednisolone was associated with lower mortality in older children.

Our mortality contrasts with previous reports, which have found lower pediatric COVID-19 mortality. Most large studies have been from HICs and report mortality rates ranging from 1.6% to 6.5%, with most deaths occurring in older children with comorbidities ^4–7 20 21^. However, the low mortality for critical pediatric COVID-19 in HICs might not translate to LMICs. Our findings may reflect the variability in pediatric COVID-19 mortality between HICs and LMICs described by Kitano ^12^. Some Brazilian studies found low mortality but had many subjects from private hospitals with a socioeconomic profile similar to HIC populations ^8 9^. Most other studies from LMICs demonstrate higher mortality, with younger age identified as a mortality risk ^10 22 23^. There is likely a multifactorial explanation for the higher mortality we observed. Our cohort included many subjects from LMICs, and our restrictive eligibility criteria likely resulted in a more critically ill cohort. Other large studies often included subjects who were not critically ill or had lower rates of diagnoses or treatments associated with higher mortality (e.g. ARDS, IMV, vasopressors) ^4 6 7^. Our cohort may better reflect the pediatric illness spectrum and severity seen with emergence of the delta variant of SARS-CoV2.

Our results also suggest that there may be distinct age-related phenotypes in pediatric critical COVID-19. Younger children more commonly had respiratory disease, hypoxemic respiratory failure, and secondary infections such as bacteremia and pneumonia, all of which might confer a higher risk of death. This contrasts with older children, who more frequently had gastrointestinal or cutaneous symptoms, developed MIS-C, and received treatments associated with lower mortality. While the impact of age on outcomes appears dramatic in our data, prior studies have had conflicting findings related to age and illness severity ^24 25^. However, these studies evaluated children in HICs and had lower overall mortality. The age-related mortality differences we observed are consistent with data from LMICs, and these potential regional differences deserve further investigation to understand the causative factors ^12 23^. Age-related differences included a lack of association between some treatments (IVIG, methylprednisolone, and prophylactic anticoagulation) or diagnoses (MIS-C) and improved mortality in younger children as well as a larger relative impact for acute kidney or liver injury compared to older children. The higher mortality seen in young children, even when they had MIS-C or received treatments associated with better outcomes, is concerning and warrants further investigation.

Many factors could explain the association between younger age and mortality. We saw a higher IMV rate in young children, both compared to older children in our study and to other studies ^4 6^. Frequent respiratory symptoms and hypoxemia combined with early COVID-19 guidelines recommending intubation rather than NIV may have prompted intubation early during hospitalization. IMV was associated with mortality and could also lead to iatrogenic complications. In addition, intubation itself, especially when performed in challenging circumstances (e.g., COVID-19 patient isolation or unfamiliar equipment), could lead to complications such as cardiac arrest. We previously described this, and now report that over half of young children treated with IMV on day 1 also suffered cardiac arrest ^15^. We also detected higher malnutrition rates in younger children. Research is ongoing to better understand the impact of age and patient factors on outcomes.

The impact of specific treatments on outcomes also warrants discussion. While the Recovery trial demonstrated improved outcomes for adults requiring IMV that received dexamethasone, pediatric data to support this treatment are lacking ^26^. We did not find an association between dexamethasone and survival, highlighting the challenge of extrapolating from adult trials to pediatric practice. Several therapies, most of which are frequently used for MIS-C, were associated with lower mortality, including methylprednisolone, IVIG, and prophylactic anticoagulation. Given the association between MIS-C and lower mortality, it is possible that the impact of these medications may be due to MIS-C rather than the therapies themselves. With mixed or absent data supporting many MIS-C and other COVID-19 treatments, it is important to study therapies for children in a prospective and multicenter fashion ^27 28^.

Our findings have several implications, particularly regarding LMICs and age-related outcomes. Current literature has not sufficiently evaluated the impact of critical pediatric COVID-19 in LMICs or the respiratory disease more commonly seen in young children. There has been more attention on MIS-C, a novel condition but one associated with very low mortality, rather than the COVID-19 related illnesses that we and others have found to have higher mortality^12^. This higher mortality warrants further investigation to determine whether care and outcomes can be improved, particularly in resource-limited settings. These efforts are urgent since younger children are unlikely to be vaccinated soon (if ever) due to lower population-based mortality risk and the unclear balance between vaccine benefit and risk ^29^. Vaccine availability and distribution are also limited in many LMICs compared to HICs. Finally, since some experts believe that COVID-19 will become a seasonal infection rather than being eradicated, studies are needed to determine the optimal treatment for critical COVID-19 in children, not just for adults ^30^.

This study has several strengths. First, to our knowledge, our prospective study is the largest to involve both HICs and LMICs from across Europe, North America, and Latin America. We achieved significant representation from “COVID-19 hotspots” in Latin America, a region largely ignored in prior pediatric studies of COVID-19. Our relatively large sample size, broad geographic representation, and high illness severity allowed us to evaluate mortality in a way that has not been feasible in prior studies of critical pediatric COVID-19. We also included PICUs from diverse contexts, which may better allow generalizability to pediatric critical illness due to COVID-19 at a global scale. The use of an established data tool (the ISARIC report form) and study recruitment in established regional PICU networks such as LARed Network and the Pediatric Acute Lung and Sepsis Investigators (PALISI) network allowed us to make international comparisons, provided consistency to data collection, and may allow for future comparison with other studies using the same data tools.

Our study does have several limitations. As an observational study, we are unable to determine causality between treatments and outcomes. Unrecognized factors might be responsible for the higher mortality observed with certain age groups, clinical presentations, or treatments. Further, while we had a relatively large sample size and high number of deaths compared to other pediatric studies, mortality was still infrequent, limiting our ability to account for confounders. Although we had sites from many countries, our study involved a minority of PICUs in most countries, which may limit generalizability. Even with this limitation, our study involved a wider range of countries than other studies of critical pediatric COVID-19. This allowed us to provide a perspective on populations that have been underrepresented in prior research. We did not account for regional variation in patients, clinical practice, and outcomes but acknowledge that this variability likely impacts outcomes. We are currently gathering data to study this in our cohort. Since our study ended in December 2020, extrapolations of our findings to the current pandemic with viral variants and rising vaccination rates may be challenging. However, the clinical presentations and severity seen in our cohort seem to be similar to what has been anecdotally noted with the rise of the delta variant of SARS-CoV2. Lastly, we do not have data following discharge to capture late mortality or delayed complications such as long COVID. Future studies are needed evaluate long term outcomes after pediatric critical COVID-19.

## CONCLUSIONS

This large, prospective, international cohort study of critically ill children with COVID-19 identified distinct age-related variations in disease phenotypes and mortality. A respiratory phenotype, more common in younger children, was associated with higher mortality. An inflammatory or cardiovascular phenotype (commonly MIS-C) was more common in older children and associated with lower mortality. We identified a higher overall mortality rate than what has previously been seen in critical pediatric COVID-19. The higher mortality, especially among younger children, has implications for public health and vaccination strategies in LMICs and should prompt continued pediatric-specific research examining risks for mortality and determining the best treatments for critical pediatric COVID-19.

## Supporting information

CAKE Collaborators

## Data Availability

Data are available upon reasonable request.

## Abbreviations

COVID-19: coronavirus disease 2019
ICU: intensive care unit
PICU: pediatric intensive care unit
HIC: high-income country
LMIC: low-middle-income country
MIS-C: multisystem inflammatory syndrome in children
HFNC: high flow nasal cannula
NIV: noninvasive ventilation
IMV: invasive mechanical ventilation
CRRT: continuous renal replacement therapy
ARDS: acute respiratory distress syndrome
CDC: Centers for Disease Control and Prevention
LOS: length of stay
PRISM III: Pediatric Risk of Mortality III
PELOD-2: Pediatric Logistic Organ Dysfunction 2
IQR: interquartile range
CI: confidence interval
BNP: brain natriuretic peptide
IVIG: intravenous immune globulin
ECMO: extracorporeal membrane oxygenation
HCQ: hydroxychloroquine
OR: odds ratio
aOR: adjusted odds ratio

## Acknowledgments

We are thankful to all researchers direct or indirectly involved in the CAKE Study, who made their greatest efforts to do research with courage while taking care of critically ill children and even adults with COVID-19 during the difficult times and recurrent surges and waves of the pandemic. We also are grateful to the investigators of the International Severe Acute Respiratory and emerging Infection Consortium (ISARIC) for sharing their data collection tools.

## References

1. Wang JG, Zhong ZJ, Mo YF, et al. Epidemiological features of coronavirus disease 2019 in children: a meta-analysis. Eur Rev Med Pharmacol Sci 2021;25(2):1146–57. doi: 10.26355/eurrev_202101_24685 [published Online First: 2021/02/13]

2. Swann OV, Holden KA, Turtle L, et al. Clinical characteristics of children and young people admitted to hospital with covid-19 in United Kingdom: prospective multicentre observational cohort study. BMJ 2020;370:m3249. doi: 10.1136/bmj.m3249 [published Online First: 2020/09/23]

3. Kim L, Whitaker M, O’Halloran A, et al. Hospitalization Rates and Characteristics of Children Aged <18 Years Hospitalized with Laboratory-Confirmed COVID-19 - COVID-NET, 14 States, March 1-July 25, 2020. MMWR Morb Mortal Wkly Rep 2020;69(32):1081–88. doi: 10.15585/mmwr.mm6932e3 [published Online First: 2020/08/14]

4. Feldstein LR, Tenforde MW, Friedman KG, et al. Characteristics and Outcomes of US Children and Adolescents With Multisystem Inflammatory Syndrome in Children (MIS-C) Compared With Severe Acute COVID-19. JAMA 2021;325(11):1074–87. doi: 10.1001/jama.2021.2091 [published Online First: 2021/02/25]

5. Garcia-Salido A, de Carlos Vicente JC, Belda Hofheinz S, et al. Severe manifestations of SARS-CoV-2 in children and adolescents: from COVID-19 pneumonia to multisystem inflammatory syndrome: a multicentre study in pediatric intensive care units in Spain. Crit Care 2020;24(1):666. doi: 10.1186/s13054-020-03332-4 [published Online First: 2020/11/28]

6. Tripathi S, Gist KM, Bjornstad EC, et al. Coronavirus Disease 2019-Associated PICU Admissions: A Report From the Society of Critical Care Medicine Discovery Network Viral Infection and Respiratory Illness Universal Study Registry. Pediatr Crit Care Med 2021 doi: 10.1097/PCC.0000000000002760 [published Online First: 2021/05/10]

7. Bhalala US, Gist KM, Tripathi S, et al. Characterization and Outcomes of Hospitalized Children With Coronavirus Disease 2019: A Report From a Multicenter, Viral Infection and Respiratory Illness Universal Study (Coronavirus Disease 2019) Registry. Crit Care Med 2021 doi: 10.1097/CCM.0000000000005232 [published Online First: 2021/08/14]

8. Lima-Setta F, Magalhaes-Barbosa MC, Rodrigues-Santos G, et al. Multisystem inflammatory syndrome in children (MIS-C) during SARS-CoV-2 pandemic in Brazil: a multicenter, prospective cohort study. J Pediatr (Rio J) 2020 doi: 10.1016/j.jped.2020.10.008 [published Online First: 2020/11/14]

9. Prata-Barbosa A, Lima-Setta F, Santos GRD, et al. Pediatric patients with COVID-19 admitted to intensive care units in Brazil: a prospective multicenter study. J Pediatr (Rio J) 2020;96(5):582–92. doi: 10.1016/j.jped.2020.07.002 [published Online First: 2020/08/12]

10. Nino-Taravilla C, Otaola-Arca H, Lara-Aguilera N, et al. Multisystem Inflammatory Syndrome in Children, Chile, May-August 2020. Emerg Infect Dis 2021;27(5):1457–61. doi: 10.3201/eid2705.204591 [published Online First: 2021/03/25]

11. Bhopal SS, Bagaria J, Olabi B, et al. Children and young people remain at low risk of COVID-19 mortality. Lancet Child Adolesc Health 2021;5(5):e12–e13. doi: 10.1016/S2352-4642(21)00066-3 [published Online First: 2021/03/14]

12. Kitano T, Kitano M, Krueger C, et al. The differential impact of pediatric COVID-19 between high-income countries and low-and middle-income countries: A systematic review of fatality and ICU admission in children worldwide. PLoS One 2021;16(1):e0246326. doi: 10.1371/journal.pone.0246326 [published Online First: 2021/01/30]

13. Zimmermann P, Curtis N. Why is COVID-19 less severe in children? A review of the proposed mechanisms underlying the age-related difference in severity of SARS-CoV-2 infections. Arch Dis Child 2020 doi: 10.1136/archdischild-2020-320338 [published Online First: 2020/12/03]

14. Pierce CA, Preston-Hurlburt P, Dai Y, et al. Immune responses to SARS-CoV-2 infection in hospitalized pediatric and adult patients. Sci Transl Med 2020;12(564) doi: 10.1126/scitranslmed.abd5487 [published Online First: 2020/09/23]

15. Gonzalez-Dambrauskas S, Vasquez-Hoyos P, Camporesi A, et al. Pediatric Critical Care and COVID-19. Pediatrics 2020;146(3) doi: 10.1542/peds.2020-1766 [published Online First: 2020/06/11]

16. Khemani RG, Smith LS, Zimmerman JJ, et al. Pediatric acute respiratory distress syndrome: definition, incidence, and epidemiology: proceedings from the Pediatric Acute Lung Injury Consensus Conference. Pediatr Crit Care Med 2015;16(5 Suppl 1):S23–40. doi: 10.1097/PCC.0000000000000432 [published Online First: 2015/06/04]

17. Prevention CfDCa. Information for Healthcare Providers about Multisystem Inflammatory Syndrome in Children (MIS-C) 2020 [Available from: https://www.cdc.gov/mis-c/hcp/ accessed May 13, 2021.

18. Pollack MM, Patel KM, Ruttimann UE. PRISM III: an updated Pediatric Risk of Mortality score. Crit Care Med 1996;24(5):743–52. doi: 10.1097/00003246-199605000-00004 [published Online First: 1996/05/01]

19. Leteurtre S, Duhamel A, Salleron J, et al. PELOD-2: an update of the PEdiatric logistic organ dysfunction score. Crit Care Med 2013;41(7):1761–73. doi: 10.1097/CCM.0b013e31828a2bbd [published Online First: 2013/05/21]

20. Sachdeva R, Rice TB, Reisner B, et al. The Impact of Coronavirus Disease 2019 Pandemic on U.S. and Canadian PICUs. Pediatr Crit Care Med 2020;21(9):e643–e50. doi: 10.1097/PCC.0000000000002510 [published Online First: 2020/07/11]

21. Kanthimathinathan HK, Buckley H, Lamming C, et al. Characteristics of Severe Acute Respiratory Syndrome Coronavirus-2 Infection and Comparison With Influenza in Children Admitted to U.K. PICUs. Crit Care Explor 2021;3(3):e0362. doi: 10.1097/CCE.0000000000000362 [published Online First: 2021/04/01]

22. de Farias ECF, Pedro Piva J, de Mello M, et al. Multisystem Inflammatory Syndrome Associated With Coronavirus Disease in Children: A Multi-centered Study in Belem, Para, Brazil. Pediatr Infect Dis J 2020;39(11):e374–e76. doi: 10.1097/INF.0000000000002865 [published Online First: 2020/08/23]

23. Coronado Munoz A, Tasayco J, Morales W, et al. High incidence of stroke and mortality in pediatric critical care patients with COVID-19 in Peru. Pediatr Res 2021 doi: 10.1038/s41390-021-01547-x [published Online First: 2021/05/05]

24. Graff K, Smith C, Silveira L, et al. Risk Factors for Severe COVID-19 in Children. Pediatr Infect Dis J 2021;40(4):e137–e45. doi: 10.1097/INF.0000000000003043 [published Online First: 2021/02/05]

25. Ouldali N, Yang DD, Madhi F, et al. Factors Associated With Severe SARS-CoV-2 Infection. Pediatrics 2021;147(3) doi: 10.1542/peds.2020-023432 [published Online First: 2020/12/17]

26. Group RC, Horby P, Lim WS, et al. Dexamethasone in Hospitalized Patients with Covid-19. N Engl J Med 2021;384(8):693–704. doi: 10.1056/NEJMoa2021436 [published Online First: 2020/07/18]

27. Davies P, Lillie J, Prayle A, et al. Association Between Treatments and Short-Term Biochemical Improvements and Clinical Outcomes in Post-Severe Acute Respiratory Syndrome Coronavirus-2 Inflammatory Syndrome. Pediatr Crit Care Med 2021;22(5):e285–e93. doi: 10.1097/PCC.0000000000002728 [published Online First: 2021/03/27]

28. Ouldali N, Toubiana J, Antona D, et al. Association of Intravenous Immunoglobulins Plus Methylprednisolone vs Immunoglobulins Alone With Course of Fever in Multisystem Inflammatory Syndrome in Children. JAMA 2021;325(9):855–64. doi: 10.1001/jama.2021.0694 [published Online First: 2021/02/02]

29. Obaro S. COVID-19 herd immunity by immunisation: are children in the herd? Lancet Infect Dis 2021 doi: 10.1016/S1473-3099(21)00212-7 [published Online First: 2021/04/23]

30. Mitropoulos A. COVID-19 could become a seasonal illness like the flu, experts say 2021 [updated February 12, 2021. Available from: https://abcnews.go.com/Health/covid-19-seasonal-illness-flu-experts/story?id=75830451 accessed May 13, 2021.

