## Supplementary tables for "Pediatric critical COVID-19 and mortality in a multinational cohort"

**Supplemental Tables**

**Supplemental Table 1: Number of subjects and study sites per country**

| **Country** | **Number of subjects**  **(% of total)** | **Number of Centers** |
| --- | --- | --- |
| USA | 110 (20%) | 19 |
| Canada | 2 (0.4%) | 2 |
| Puerto Rico | 2 (0.4%) | 1 |
| Ireland | 2 (0.4%) | 1 |
| Italy | 48 (8.6%) | 2 |
| Portugal | 7 (1.3%) | 1 |
| Spain | 11 (2%) | 1 |
| Turkey | 23 (4.1%) | 2 |
| Argentina | 29 (5.2%) | 8 |
| Bolivia | 2 (0.4%) | 8 |
| Chile | 39 (7%) | 11 |
| Colombia | 137 (25%) | 20 |
| Costa Rica | 17 (3.1%) | 1 |
| Honduras | 29 (5.2%) | 1 |
| Mexico | 16 (2.9%) | 1 |
| Peru | 83 (15%) | 2 |

Two sites in Uruguay and one site in Panama also screened, but no subjects were enrolled

from these countries

**Supplemental Table 2: Comparison of symptoms at presentation, including symptom category and components of each category**

| **Symptoms** | **All** | **Age < 2y**  (n=134) | **Age >2y**  (n=423) |
| --- | --- | --- | --- |
| Lower Respiratory | 262 (47%) | 75 (56%) | 187 (44%) |
| Cough | 209 (38%) | 55 (41%) | 154 (36%) |
| Wheeze | 63 (11%) | 27 (20%) | 36 (8.5%) |
| Retractions | 164 (29%) | 60 (45%) | 104 (25%) |
| Upper Respiratory | 165 (30%) | 50 (37%) | 115 (27%) |
| Sore Throat | 91 (16%) | 3 (2.2%) | 88 (21%) |
| Rhinorrhea | 95 (17%) | 49 (37%) | 46 (11%) |
| Ear Pain | 6 (1%) | 1 (0.7%) | 5 (1.2%) |
| Systemic | 469 (84%) | 91 (68%) | 378 (89%) |
| Fever | 428 (77%) | 80 (60%) | 348 (82%) |
| Fatigue | 310 (56%) | 55 (41%) | 255 (60%) |
| Gastrointestinal | 327 (59%) | 37 (28%) | 290 (69%) |
| Abdominal Pain | 241 (43%) | 14 (10%) | 227 (54%) |
| Vomiting | 238 (43%) | 24 (18%) | 214 (51%) |
| Diarrhea | 187 (34%) | 24 (18%) | 163 (39%) |
| Neurologic | 214 (38%) | 41 (31%) | 173 (41%) |
| Headache | 114 (20%) | 3 (2.2%) | 111 (26%) |
| Confusion | 107 (19%) | 32 (24%) | 75 (18%) |
| Seizure | 48 (8.6%) | 13 (9.7%) | 35 (8.3%) |
| Mucocutaneous | 208 (37%) | 21 (16%) | 187 (44%) |
| Rash | 147 (26%) | 16 (12%) | 131 (31%) |
| Conjunctivitis | 118 (21%) | 8 (6%) | 110 (26%) |
| Skin Ulcers | 8 (1.4%) | 1 (0.7%) | 7 (1.7%) |
| “Strawberry” Tongue/Oral Lesions | 64 (12%) | 6 (4.5%) | 58 (14%) |
| Red Hands/Feet | 105 (19%) | 10 (7.5%) | 95 (23%) |
| Non-Specific | 299 (54%) | 58 (43%) | 241 (57%) |
| Chest Pain | 44 (7.9%) | 0 | 44 (10%) |
| Myalgias | 131 (24%) | 9 (6.7%) | 122 (29%) |
| Arthralgias | 49 (8.8%) | 2 (1.5%) | 47 (11%) |
| Shortness of Breath | 192 (34%) | 52 (39%) | 140 (33%) |
| Lymphadenopathy, general | 53 (9.5%) | 4 (3%) | 49 (12%) |
| Lymphadenopathy, cervical | 49 (9%) | 5 (3.9%) | 44 (11%) |
| Bleeding | 14 (2.5%) | 3 (2.2%) | 11 (2.6%) |

Data are n (%)

**Supplemental Table 3: Complete list of hospital diagnoses and complications with comparison between children < 2 years and children > 2 years old**

| **Diagnosis/Complication** | **All** | **Age <2 years** | **Age >2 years** | p-value^a^ |
| --- | --- | --- | --- | --- |
| Viral Pneumonia/Pneumonitis | 183 (33%) | 60 (45%) | 123 (29%) | 0.001 |
| Bacterial Pneumonia | 113 (20%) | 26 (19%) | 87 (21%) | 0.81 |
| ARDS | 157 (28%) | 42 (31%) | 117 (27%) | 0.38 |
| Pneumothorax | 15 (2.7%) | 5 (3.7%) | 10 (2.4%) | 0.37 |
| Pleural Effusion | 79 (14%) | 9 (6.7%) | 70 (17%) | 0.004 |
| Bronchiolitis | 26 (4.7%) | 23 (17%) | 3 (0.7%) | <0.001 |
| Meningitis | 29 (5.2%) | 9 (6.7%) | 20 (4.7) | 0.37 |
| Seizure | 53 (9.5%) | 15 (11%) | 38 (9%) | 0.50 |
| Stroke | 19 (3.4%) | 5 (3.7%) | 14 (3.3%) | 0.79 |
| Heart Failure | 86 (15%) | 16 (12%) | 70 (17%) | 0.22 |
| Myocarditis/Cardiomyopathy | 101 (18%) | 11 (8.2%) | 90 (21%) | <0.001 |
| Arrhythmia | 39 (7%) | 9 (6.7%) | 30 (7.1%) | >0.99 |
| Cardiac Arrest | 56 (10%) | 23 (17%) | 33 (7.8%) | 0.003 |
| Bacteremia | 68 (12%) | 28 (21%) | 40 (9.5%) | <0.001 |
| Coagulopathy | 98 (18%) | 18 (13%) | 80 (19%) | 0.15 |
| Anemia | 198 (36%) | 57 (43%) | 141 (33%) | 0.06 |
| Rhabdomyolysis | 6 (1.1%) | 1 (0.7%) | 5 (1.2%) | >0.99 |
| Acute Kidney Injury | 88 (16%) | 13 (9.7%) | 75 (18%) | 0.03 |
| Gastroenteritis | 11 (2%) | 2 (1.5%) | 9 (2.1%) | >0.99 |
| Pancreatitis | 13 (2.3%) | 1 (0.7%) | 12 (2.8%) | 0.32 |
| Acute Hepatic Injury | 51 (9.2%) | 6 (4.5%) | 45 (11%) | 0.04 |
| MIS-C | 188 (34%) | 20 (15%) | 168 (40%) | <0.001 |

Data are n (%)

^a^Comparisons using Fisher’s exact test

Abbreviations: ARDS = acute respiratory distress syndrome; MIS-C = multisystem inflammatory syndrome in children

**Supplemental Table 4: Univariate associations with mortality for variables included in multivariable analysis using logistic regression analysis**

| **Variable** | **OR (95% CI)** | **Variable** | **OR (95% CI)** |
| --- | --- | --- | --- |
| **Comorbidity** |  | **Medications** |  |
| Any Comorbidity | 3.06 (1.42, 6.60) | Prophylactic Anticoagulation | 0.42 (0.21, 0.84) |
| Cardiac Comorbidity | 3.28 (1.40, 7.11) | Dexamethasone | 1.13 (0.62, 2.07) |
| Pulm. Comorbidity | 5.32 (2.03, 13.9) | Methylprednisolone | 0.39 (0.20, 0.75) |
| Cancer | 3.06 (1.42, 6.60) | IVIG | 0.31 (0.16, 0.58) |
| Obesity | 0.47 (0.16, 1.34) | **Diagnoses/Complications** |  |
| Malnutrition | 3.94 (1.79, 8.67) | Viral Pneumonia | 2.04 (1.17, 3.58) |
| **Clinical Findings** |  | Bacterial Pneumonia | 2.88 (1.61, 5.16) |
| Admission O_2_ Saturation <94% | 2.50 (1.39, 4.50) | ARDS | 6.86 (3.77, 12.5) |
| Lower Respiratory | 3.47 (1.87, 6.43) | Seizures | 3.12 (1.52, 6.37) |
| Gastrointestinal | 0.46 (0.26, 0.80) | Myocarditis | 0.72 (0.33, 1.58) |
| Mucocutaneous | 0.33 (0.16, 0.67) | Bacteremia | 6.54 (3.52, 12.1) |
| **Day 1 Support** |  | Acute Kidney Injury | 5.26 (2.91, 9.52) |
| IMV | 3.67 (2.06, 6.41) | Acute Liver Injury | 2.79 (1.34. 5.82) |
| Vasopressors | 2.06 (1.18, 3.59) | MIS-C | 0.21 (0.09, 0.50) |

Abbreviations: Pulm. = Pulmonary; IMV = invasive mechanical ventilation; IVIG = intravenous immune globulin;

ARDS = acute respiratory distress syndrome; MIS-C = multisystem inflammatory syndrome in children
