## Supplementary material for "Pediatric critical COVID-19 and mortality in a multinational cohort": CAKE Collaborators

**Study Sites and Collaborators:**

Akron Children's Hospital, Akron, OH, USA (Ryan Nofziger, MD)

Albany Medical Center, Albany, NY, USA (Shashikanth Ambati MBBS)

Ankara University, Ankara,Turkey (Anar Gurbanov, MD)

Arkansas Children's Hospital, Little Rock, AR, USA (Ronald Sanders, MD)

Brown University/Hasbro Children’s Hospital, Providence, RI,USA (Lee Polikoff, MD)

Children's Health Ireland, Dublin,Ireland (Siobhan Whelan, MBBS)

Children's Hospital Vittore Buzzi, Milan, Italy (Anna Camporesi, MD)

CHU de Quebec, Quebec City,Quebec, Canada (Conall Francoeur, MD)

Clínica Alemana de Santiago, Santiago, Chile (Francisca Castro, MD)

Clínica el Rosario, Universidad de Antioquia, Medellín, Colombia (Claudia Beltrán, MD)

Clínica Infantil Colsubsidio, Bogota, Colombia (Rosalba Pardo, MD)

Clínica Infantil Santa María del Lago, Bogota, Colombia (Gonzalo Vega, MD)

Clínica Las Condes, Santiago, Chile (Mauricio Yunge, MD)

Clínica Universitaria Colombia, Bogota, Colombia (Lorena Acevedo, MD)

Clínica UROS, Neiva, Colombia (Ivan Jose Ardila, MD)

Complejo Asistencial Víctor Ríos Ruiz, Los Angeles, Chile (Diego Aranguiz, MD)

Dell Children's Hospital, Austin, TX, USA (Samantha Dallefeld, MD)

Fundación Cardioinfantil-Instituto de Cardiología, Bogota, Colombia (Martha I Alvarez-Olmos, MD, MPH, Universidad del Bosque and Jaime Fernandez-Sarmiento, MD, PhD(c), Universidad de la Sabana)

Fundación Clínica Infantil Club Noel, Cali, Colombia (Arieth Figueroa Vargas, MD and Maribel Valencia Benavides, MD)

Fundación HOMI, Bogota, Colombia (Juan David Roa, MD, MSc)

Fundación Valle del Lili, Cali, Colombia (Rubén Lasso Palomino, MD)

Gaslini Children’s Hospital, Genoa, Italy (Alessia Franceschi, MD)

Hospital Carlos Van Buren, Valparaiso. Chile (Carina Venthur, MD)

Hospital Casa de Galicia, Montevideo, Uruguay (Sebastian Gonzalez-Dambrauskas, MD)

Hospital Clínico La Florida, Dra Eloísa Díaz Insunza, Santiago, Chile (Camila Ampuero, MD)

Hospital Daniel Bracamonte, Potosi, Bolivia (Jhovana E. Paco Barral, MD)

Hospital de Emergencias Villa El Salvador, Lima, Peru (Jaime Tasayco Muñoz, MD and Jesús Domínguez Rojaz, MD)

Hospital de Niños Dr. Mario Ortiz Suarez, Santa Cruz de la Sierra, Bolivia (Francisca Rafael Patricio, MD)

Hospital de Pediatría Juan P. Garrahan, Buenos Aires, Argentina (Solana Pellegrini, MD, Marcela Zuazaga, MD and Silvana Brusca, MD)

Hospital de Santa Maria, Lisbon, Portugal (Marisa Viera, MD)

Hospital del Niño Dr. Ovidio Aliaga Uría, La Paz, Bolivia (Vladmir Ivan Aguilera Avendano, MD)

Hospital del Niño Manuel Ascencio Villarroel, Cochambamba, Bolivia (Alejandro F. Martínez L., MD and Thelma E. Terán M., MD)

Hospital del Niño Sor Teresa Huarte Tama, Sucre, Bolivia (Mariela Coronado Lujan, MD)

Hospital Dr. Exequiel González Cortés, Santiago, Chile (Fabiola Castro Mancilla, MD)

Hospital El Carmen de Maipú, Santiago, Chile (Franco Diaz-Rubio, MD)

Hospital El Cruce, Buenos Aries, Argentina (Karina Cinquegrana, MD and Alicia Sandoval, MD)

Hospital Félix Bulnes Cerda, Santiago, Chile (Andrea Gonzalez, MD and Marta Zamora, MD)

Hospital General de Medellín, Medellin, Colombia (Yurika Lopez-Alarcon, MD)

Hospital Gregorio Marañón, Madrid, Spain (María Slöcker Barrio, MD and Javier Urbano Villaescusa, MD, PhD)

Hospital Hernán Messuti Rivera, Cobija, Bolivia (Humberto Camacho Delgadillo, MD)

Hospital Infantil Los Ángeles, Pasto, Colombia (Liliana Mazzillo Vega, MD)

Hospital Infantil Rafael Henao Toro, Manizales, Colombia (Beatriz Giraldo, MD)

Hospital Materno Infantil Boliviano Japonés, Trinidad, Bolivia (Pitas Suarez, MD and Miguel Cespades Lesczinsky, MD)

Hospital Materno Infantil José Domingo De Obaldía, David, Panama (Jorge Omar Castillo, MD)

Hospital Municipal de Agudos Leonidas Lucero, Bahía Blanca, Argentina (Juan Pablo Fabris, MD and Carolina Paladino, MD)

Hospital Nacional de Niños Dr. Carlos Saenz Herrera,CCSS, San Jose, Costa Rica (Silvia Sanabria, MD, Erika Urena-Chavarría, MD and Adriana Yock-Corrales, MD, MSc)

Hospital Nacional Edgardo Rebagliati Martins, Lima, Peru (Gaudi Quispe Flores, MD and Manuel Munaico Abanto, MD)

Hospital Nacional Profesor A. Posadas, Buenos Aires, Argentina (Miriam Colombo, MD and Ana Carola Blanco, MD)

Hospital Pablo Tobón Uribe, Medellin, Colombia (Byron Enrique Pineres-Olave, MD)

Hospital Padre Hurtado, Santiago, Chile (Ricardo Carvajal Veas, MD)

Hospital Pediátrico Alexander Fleming, Mendoza, Argentina (Patricia Correa, MD)

Hospital Pediátrico Universitario, San Juan, Puerto Rico (Ricardo Garcia-De Jesus, MD)

Hospital Policial, Montevideo, Uruguay (Arani Ferre, MD)

Hospital Regional de Antofagasta, Antofagasta, Chile (Pietro Pietroboni, MD)

Hospital Regional del Norte IHSS, San Pedro Sula, Honduras (Edwin Mauricio Cantillano, MD and Linda Banegas Pineda, MD)

Hospital Regional San Juan de Dios, Tarija, Bolivia (Nils Casson Rodriguez, MD)

Hospital Sotero del Rio, Santiago, Chile (Agustin Cavagnaro, MD and Adriana Wegner, MD)

Hospital Susana López de Valencia, Popayan, Colombia (Eliana Zemanate, MD and Emilce Beltran Zuñiga, MD)

Hospital Universitario San Ignacio, Bogota, Colombia (Maria Alejandra Suarez, MD and Deyanira Quiñonez, MD)

Hospital Universitario San Jorge, Pereira, Colombia (Leonardo Valero, MD)

Hospital Zonal de Trelew, Trelew, Argentina (Alejandra Repetur, MD)

Hospital de Niños Sor María Ludovica, La Plata, Argentina (Pablo Castellani, MD and Adriana Bordogna, MD)

IMAT Oncomedica, Montería, Colombia (Alfredo De la Hoz Pastor, MD)

Instituto Roosevelt, Bogota, Colombia (Evelyn Obando Belalcazar, MD)

John R. Oshei Children's Hospital of Buffalo, Buffalo, NY, USA (Andrew Prout, MD)

Juan Pablo II Hospital, Corrientes, Argentina (Roberto Jabornisky, MD)

Lucile Packard Children's Hospital, Palo Alto, CA, USA (Andy Wen, MD)

Lurie Children's Hospital, Chicago, IL, USA (Bria Coates, MD)

Medical College of Georgia, Augusta, GA, USA (Christopher Watson, MD, MPH)

Medical University of South Carolina, Charleston, SC, USA (Elizabeth Mack, MD, MS)

Medstar Georgetown, Washington, DC, USA (Jahee Hong, MD)

Nationwide Children's Hospital, Columbus, Ohio, USA (Todd Karsies, MD, MPH)

New York-Presbyterian/Weill Cornell Medicine, New York, NY, USA (Steven Pon, MD)

NYU Langone, New York City, NY, USA (Heda Dapul, MD)

Rainbow Babies and Children's Hospital, Cleveland, Ohio, USA (Steven Shein, MD)

Saglik Bilimleri University Gazi Yasargil Training and Research Hospital, Istanbul, Turkey (Murat Kangin, MD)

Sociedad de Cirugía Hospital de San José, FUCS, Bogota, Colombia (Pablo Vasquez-Hoyos, MD, MSc)

St. Barnabas Medical Center, Livingston, NJ, USA (Shira Gertz, MD)

Ste. Justine, Montreal, QC, Canada (Laurence Ducharme-Crevier, MD)

Stony Brook Children's Hospital, Stony Brook, NY, USA (Ilana Harwayne-Gidansky, MD)

UMAE Hospital de Pediatría CMN Siglo XXI “Dr. Silvestre Frenk Freund”, IMSS Mexico City, Mexico (Marisol Fonseca-Flores, MD and Juan Carlos Nunez-Enriquez, PhD)

UMHES Santa Clara de la Subred Centro Oriente, Bogota, Colombia (Armando Leon Villanueva, MD and Ledys Maria Izquierdo Borrero, MD)

University of Oklahoma Health Sciences Center, Oklahoma City, OK, USA (Teddy Muisyo, MD)

University of Virginia Medical Center, Charlottesville, VA, USA (Michael Spaeder, MD, MS)
